# A Hybrid Simulation Model of HIV Program Interventions: From Transmission Behavior to Macroeconomic Impacts

**DOI:** 10.1101/2024.06.06.24308576

**Authors:** William Crown, Erin Britton, Moaven Razavi, Yiqun Luan, Senthil Veerunaidu, Jennifer Kates, Gary Gaumer, Monica Jordan, Clare L. Hurley, A.K. Nandakumar

## Abstract

While several studies have examined the public health effects of HIV investments, there is a smaller literature that examines the macroeconomic impacts of such investments. The most recent efforts have utilized econometric analyses of country-level panel data, as well as demographic simulations of HIV spending. However, there are no studies that examine the linkages between the behavioral responses to public health investments, demographic changes, and, ultimately, macroeconomic performance. This study demonstrates the feasibility of such a linkage. We develop an agent-based simulation model (ABM) of a Peer Navigator Program to support engagement in HIV therapy in Tanzania. Results from the ABM are weighted to reflect the Tanzania population and fed into the SPECTRUM model. This generates detailed demographic forecasts that are translated into macroeconomic impacts using labor force participation rates from the International Labor Organization, along with an econometric model of gross domestic product (GDP). Although the simulated effects of the Peer Navigator Program in Tanzania are small, the paper demonstrates the feasibility of linking behavioral ABM simulations of program impacts to subsequent demographic effects and, finally, macroeconomic performance.

## Introduction

Several studies have demonstrated that large public health investments to address HIV in low- and middle-income countries have had positive impacts in reducing mortality (Gaumer G et al., 2024). Mortality reductions directly influence the size of the labor force with subsequent implications for the productive capacity of economies. Despite the size of the positive impacts of public investment in HIV programs on population health, the literature estimating the macroeconomic benefits of such investment is relatively small (Bloom D et al., 2018; Crown W et al., 2023; Economist Impact, 2023; Piabuo S et al., 2017; Remes J et al., 2020).

In addition to the direct impact of mortality reductions on labor force size and the productive capacity of economies, economic growth, in the long run, also enhances the ability of a society to invest in further educational and health care infrastructure creating a positive feedback loop stemming from the original HIV program investments (Piabuo S et al., 2017). Finally, it is plausible to expect that any large-scale public program to address HIV would have direct and indirect demand-side macroeconomic impacts as spending from such programs work their way through countries’ economies over time. Specifically, investments in healthcare infrastructure generate income for health care workers which is then spent creating subsequent income for people working in other sectors. This income is then re-spent and, via Keynesian macroeconomic multipliers, generates potential benefits worth multiples of the original expenditure.

A large literature has demonstrated that investments in human capital and health are associated with economic growth (Bloom D et al., 2018; Bloom DE et al., 2020; Collin M et al., 2018; Piabuo S et al., 2017; Remes J et al., 2020; Vogl TS, 2012; World Bank, 2020). However, analysis of the economic and educational impacts of health investments made by specific programs is less common. One example is our set of recent analyses done to assess the association between PEPFAR, the U.S. government’s global HIV program and largest commitment by any country addressing a single disease (Bor J et al., 2012; Guo Y et al., 2012; Resch S et al., 2011; Thirumurthy H et al., 2012) and several health and non-health-outcomes. We found that PEPFAR investments were associated with large, significant reductions in all-cause mortality (Gaumer G et al., 2024), reductions in maternal and child mortality and increases in childhood immunizations (Gaumer et al., 2023), as well as significant, positive, impacts on educational levels and gross domestic product (GDP) (Crown W et al., 2023). However, despite these impacts on mortality, educational levels, and GDP we did not find positive impacts on employment. Indeed, descriptive analyses of employment levels over the pre/post PEPFAR periods indicate that employment levels remained remarkably flat. In contrast, some previous studies that have found positive employment effects associated with HIV public health programs have used person and household-level data (Wagner Z et al., 2015). This suggests that within-country and cross-country variability in employment effects may be masked by such aggregate trend data. A recent study by the Economist Global Impact Team found substantial positive macroeconomic effects of HIV investments (Economist Impact, 2023).

### Why simulation methods are needed

Health care delivery systems are complex and often highly fragmented. They are social systems consisting of many different groups or agents including governments, payers, and providers responsible for delivering health care services to patients (Marshall D. et al., 2015). Social systems are inherently complex because each of the agents in these systems makes decisions, interacts among themselves, and interacts with other parts of the system in an interdependent nature. The complexity of such interactions leads to emergent behavior that is difficult to predict. Although modeling approaches such as decision trees and Markov models have been widely used as methods to evaluate health care interventions, these approaches may not be sufficient for analyzing complex health care delivery systems. Instead, dynamic simulation modeling offers advantages, given recent advances in accessible computing power and data analytics that make it possible to simulate the impact of system interventions on health care delivery systems without costly and time-consuming direct experimentation.

The results of such simulation models can anticipate the comparative effectiveness of a novel system intervention as well as its cost-effectiveness. Three dynamic simulation modeling methods are well suited for and commonly applied to these types of problems: system dynamics (SD), discrete event simulation (DES), and agent-based modeling (ABM). (Crane GJ et al., 2013; Homer JB et al., 2006; Liu P et al., 2014; Seila AF et al., 2009). For this analysis, we used ABM, as described below.

### Agent-based simulation models

ABM is a simulation method for modeling dynamic, adaptive, and autonomous systems. (Marshall D. et al., 2015) At the heart of an ABM model, there are autonomous and interacting objects, who have the ability to react to the changes in their environment and actively make decisions. These decision units are called agents. In healthcare contexts the most notable agents are patients, households, and healthcare providers who interact with one another. Each agent exists within an environment and their next actions are based on the current state of their environment. An agent senses its environment and behaves accordingly on the basis of defined decision rules. The three core concepts that form the basis for an ABM are agency, dynamics, and structure. Dynamics means that both the agents and their environment can change, develop, or evolve over time. Structure is emergent from agent interaction. How human populations will tend to congregate and interact in certain locations such as the workplace, marketplace, or school is an example of structure. Patient centered ABMs enable the researchers to simulate the health seeking behaviors of the agents in a changing environment, or in response to incentives and stimuli, along with the impacts they may have on other agents’ behaviors or even on their environment. ABM is a rapidly maturing health modeling technique that is well suited for addressing public health planning and policy needs, as well as supporting health care infrastructure investment decisions. The ability of ABMs to simulate the effects of introducing a program and changing its features is a powerful tool for assisting public health planning and program implementation.

### The need for hybrid simulation models

Simultaneous or sequential estimates of the effects of health seeking behaviors and outcomes on subsequent social and economic indicators require a hybrid cascaded modeling approach. Standalone simulation models have been widely used to model the transmission of HIV. (Abuelezam NN et al., 2013) However, the use of ABMs to model the behavioral response to HIV programs has been limited (Vermeer W et al., 2022) and there are no examples of cascading such models to other modeling tools to examine their implications for demographic and macroeconomic impacts. Any large-scale public health investment program such as PEPFAR will operate through the response of agents to its initiatives. For example, the establishment of a new HIV clinic will require individuals to travel to the clinic to get tested. Among those with HIV awareness, some must decide whether to initiate antiretroviral therapy (ART) and continue on ART to maintain their own health until they are no longer infectious. Some individuals will discontinue ART and could become sick and/or infectious once again. The point is that much of the impact of public health initiatives is the cumulative result of the behavioral responses that the program triggers in a large number of individuals or agents. The ability to model these behavioral responses and understand their impacts is key to understanding the dynamics of how a program can achieve its intended effects. Consequently, it is critically important to develop ABM simulation capabilities to assist large-scale public health programs such as PEPFAR in planning and program evaluation.

The ultimate goal of any public health investment is to reduce morbidity and mortality in the target population. Much of the research on HIV interventions has focused on estimating the effects of such interventions on patient outcomes using statistical methods from epidemiology and econometrics, as well as simulation methods. Research on the economic impacts of specific HIV programs such as PEPFAR is far more limited. The most recent, and ambitious, effort to simulate the macroeconomic impacts of HIV investments is the work of The Economist Global Impact Team. Their study links demographic estimates of the impacts of HIV spending using the SPECTRUM model to estimate demographic impacts and subsequent macroeconomic impacts (Stover J et al., 2014; Economist Impact, 2023). SPECTRUM is a sophisticated HIV policy analysis tool that incorporates the ability to simulate the demographic effects of aggregate inputs such as program spending or changes in ART or viral load suppression (VLS) at a country-specific level (Stover et al., 2014). It incorporates detailed demographic modeling dynamics based upon population structure by age and sex, birth rates, and impacts of ART and VLS on mortality by age and sex. In this analysis, we extend the work of the Economist Impact (2023) report by adding an ABM component to the model. Rather than assuming the levels of the program inputs that feed into SPECTRUM, we use an ABM to simulate them. In so doing, we are able to trace the behavioral responses to a public health intervention (ABM) on demographic dynamics (SPECTRUM) and finally on macroeconomics. We develop and test this cascaded hybrid modeling capability by simulating a peer navigator program in Tanzania.

## Methods

We developed an ABM to simulate a peer navigation program and estimate the macroeconomic impact of public health investments in HIV care continuity. This model integrated the change in adult ART retention to project population changes over a ten-year period and linked demographic data with labor force participation rates to estimate the overall GDP.

### Simulating a Peer Navigator Program in Tanzania

To pilot the utility of this hybrid modeling approach, we build on prior research demonstrating the potential for married women with high agency to positively influence their social networks through peer navigation (Sherafat-Kazemzadeh et al., 2021). The intervention in our ABM was based on recent evidence from a randomized controlled trial in Kenya which demonstrated the potential of peer navigation to improve care re-engagement and retention. In this trial, Geng et al. (2023) evaluated the effects of different strategies to influence treatment initiation and retention and found that peer navigators meaningfully improved treatment success among those with lapses in continuity.

### Tanzania: an illustration

Given these results, we evaluated the effects of a peer navigator program on demographic and macroeconomic outcomes using a hybrid approach linking an ABM, SPECTRUM, and macroeconomic model using data from Tanzania. The structure of the model is illustrated in Fig 1.

**Fig 1.**
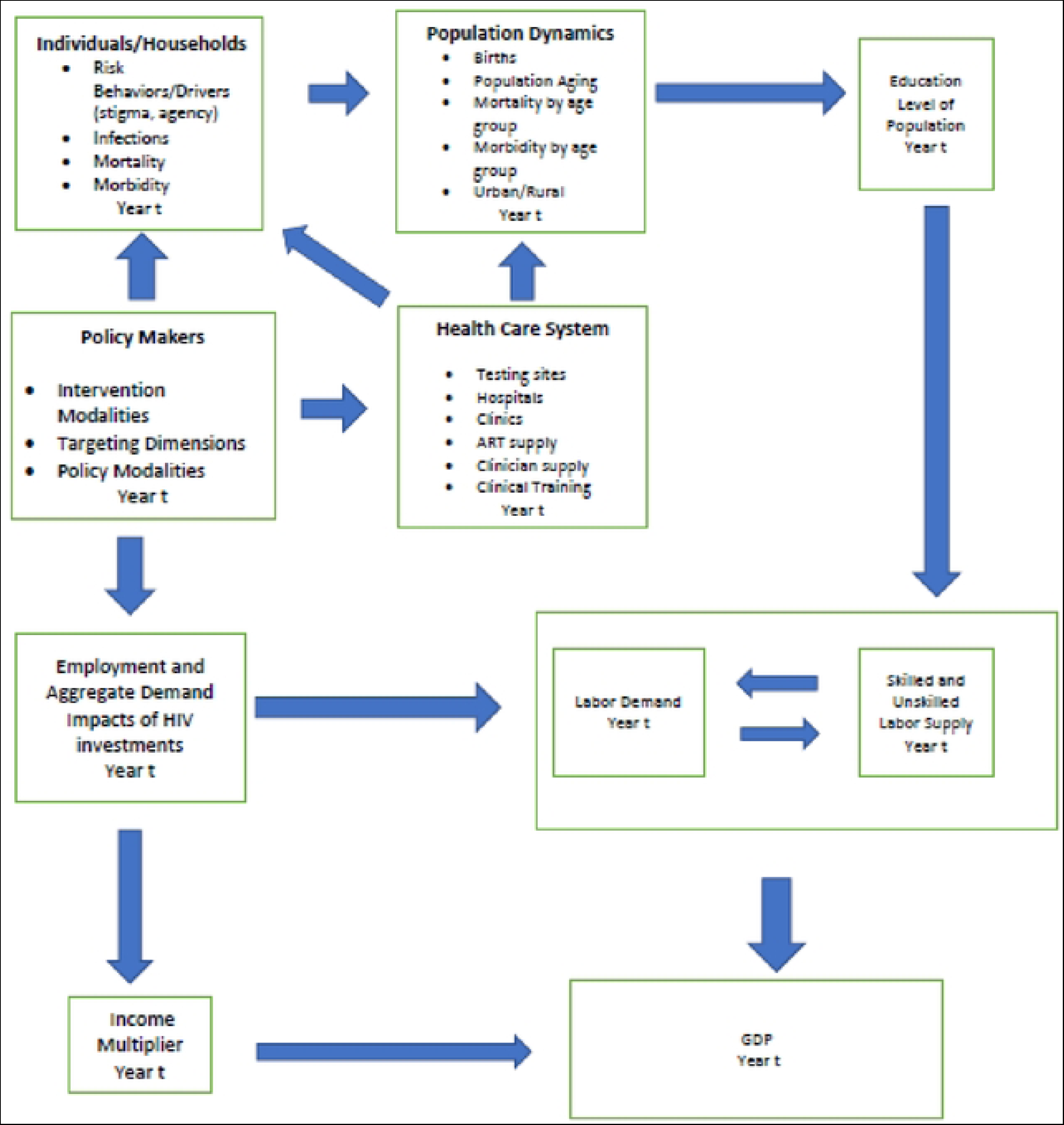
ABM and System Dynamics Model of HIV Policy, HIV Mortality, Employment, and Gross Domestic Product.

First, consider the Policy Makers component in Fig 1. Working clockwise around the figure, the model assumes that health investments in HIV programs such as PEPFAR create improvements in health care infrastructure which improves access to care, as well as behavioral incentives for households and individuals. For example, building a new clinic that supports the provision of antiretroviral therapy increases the supply of therapy for patients and households in the proximity of the new clinic. An ABM could also model the effects of clinic proximity on patient behavior by assigning probabilities that patients will come into the clinic to be tested that vary by distance. Thus, the Health Care System and Individuals/Household components are where the agent-based modeling takes place. Ultimately, the actions of agents in these two components results in changes in HIV testing and awareness, treatment with ART, and VLS. In turn, these changes in health processes and outcomes as outputs from the ABM would be expected to impact mortality, an important element of the Population Dynamics component.

### Integrating ABM with SPECTRUM

Rather than building a population dynamics component from scratch, we elected to use the existing capabilities of the SPECTRUM model. The SPECTRUM model was particularly attractive because it was specifically developed to support HIV policy analyses. The Population Dynamics component of SPECTRUM forecasts single-year population levels by sex over a 10-year forecast period using country-specific historical data on population age/sex structure, birth rates, and mortality rates. An HIV disease transmission model relates transmission of the disease and mortality to VLS. Thus, the outputs from the ABM can be entered as inputs into SPECTRUM to model their effects on the forecasted age/sex structure of the population. To link the ABM results to the SPECTRUM model, the distribution of agents used for the simulation was scaled to the “real world” population distribution of men and women in Tanzania. The weighted outputs from the ABM were then incorporated into SPECTRUM by modifying the percent of PLHIV receiving ART in the adult ART program statistics component of the AIDS Impact Model (AIM).

### Linking Population Dynamics to Economic Outcomes

Population dynamics, in turn, determine the size of the population moving through the various levels of the educational system and have a strong influence on the formation of human capital that contributes to the productivity of the labor force. Population dynamics also determine the age-specific size of the labor force. Together, these two components account for the aggregate human capital available in the labor force to produce the goods and services needed by the economy. We apply age/sex labor force participation rates from the International Labor Organization (ILO)(International Labour Organization, 2023) to the SPECTRUM demographic forecasts to generate estimates of the labor force over the forecast period. The historical and forecast values of the population and labor force size for Tanzania are shown in Fig 2. The left panel shows the historical and forecast population values while the right panel reports the historical labor force values and forecast after applying the ILO labor force participation rates to the population data.

**Fig 2.**
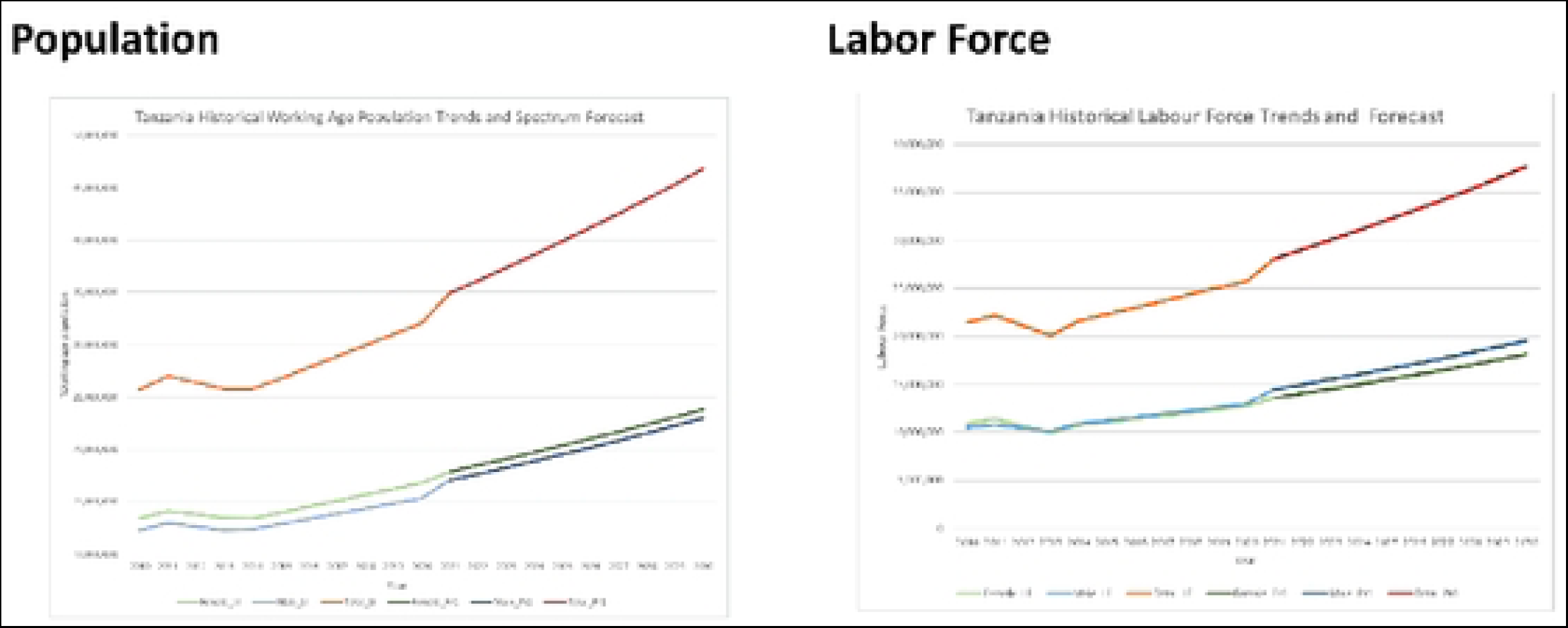
Tanzania Historical Population Trends and Baseline Spectrum Forecast and Associated Labor Force Estimates.

We then estimate an econometric model of GDP and use it to forecast GDP levels in response to changes in the productive capacity of the labor force using a similar specification to recent work in the literature (Bloom D et al., 2018; Economist Impact, 2023)

Due to limitations in the number of historical observations available for Tanzania we estimate the model using observations from 31 PEPFAR countries that prepare Country Operational Plans (COPs) and then evaluate how well the model predicts historical levels for Tanzania GDP (US Department of State, 2022). We chose PEPFAR COP countries for the modeling of GDP because these countries often have the greatest unmet HIV needs and generally receive the largest budget allocations.

The GDP model, estimated in double-log form as a Cobb-Douglas production function, explains 94 percent of the variation in the log of GDP over time. GDP is modeled as a function of the total size of the labor force, secondary school enrollment, urban population percentage, and yearly trends. The pattern of predicted and actual values when the historical observations for Tanzania are run through the model to estimate historical levels of GDP are very similar, indicating that the model appears to be well-calibrated for Tanzania even though it was estimated using data from the broader sample of COP countries. Estimation and calibration details for the GDP model are reported in the on-line supplemental materials.

### ABM for Modeling Peer Navigation in Tanzania

The objective of the ABM was to build upon our earlier work on agency of married women by simulating an intervention that targets married women to influence engagement and retention in HIV care. Agency and knowledge distributions for married women were estimated using PHIA surveys in Malawi, Tanzania, and Zambia, 2016-2017. (Sherafat-Kazemzadeh et al., 2021) We used outputs from the ABM as inputs into the SPECTRUM model to estimate population effects in Tanzania.

The ABM intervention modeled the ability of high-agency married women as peer navigators to influence family and community engagement and retention in care. Community components included variation in household size, composition, community density, and social connectedness. Employment and education were hypothesized to drive routine movement to other locations, generating social interactions. In addition, care seeking at clinics generated decision-making related to testing and treatment uptake.

In the model we simulate health seeking behavior of the agents as they make decisions dynamically about whether to migrate, seek care, adhere to their treatment plan, or return for follow-up appointments. Individuals in the model are assumed to have specific family and community networks. For example, one individual might live in a household with 3 people and have 14 social connections. Another might live in a household with 5 people and have 7 social connections. Social interactions in various settings such as school or work present opportunities to leverage the empowerment of married women to influence their household and social peers. Women who are high agency and engaged in care enter the pool of agents to be selected by the model as peer navigators.

In the scenario used for this research, 2,000 households and 10,000 married women agents participated in the ABM simulation. We also tested other model scenarios with smaller agency sizes which indicated that model results converged with approximately 1,000 households and 5,000 married women agents. However, we reported the results from the simulation with the largest agent sizes of 2,000 households and 10,000 married women. The models simulated the impacts of the peer navigator program over one, two, and three-year periods.

## Results

To pilot estimating the macroeconomic impact of public investments in HIV programs, we developed a hybrid simulation model of GDP in Tanzania linking the results of an ABM simulating the change in adult ART retention through a hypothetical peer navigation program with SPECTRUM population projections and labor force participation rates from ILO. We found that recruiting married women who reach VLS to lead a community-based peer navigation program promoting ART re-engagement increased ART participation and VLS among men and women; however, over a ten-year time horizon, modest mortality reductions had a small impact on labor force size and GDP. This indicates that changes in macro-level trends are likely to obscure important gains toward program-specific goals, particularly when interventions target small, vulnerable populations including children. Leveraging the social capital of women may be an important strategy to achieve HIV care cascade goals for men and children, but these gains may be less detectable in macroeconomic trends.

As shown in Fig 3, the ABM simulation estimated that the Peer Navigation program increased ART participation for men and women by about 12-15% with no strong trend over time. The impact on VLS, however, was cumulative and significant for both men and women. By year 3, VLS was improved by 33.9 percentage points for women and 32.6 percentage points for men.

**Fig 3.**
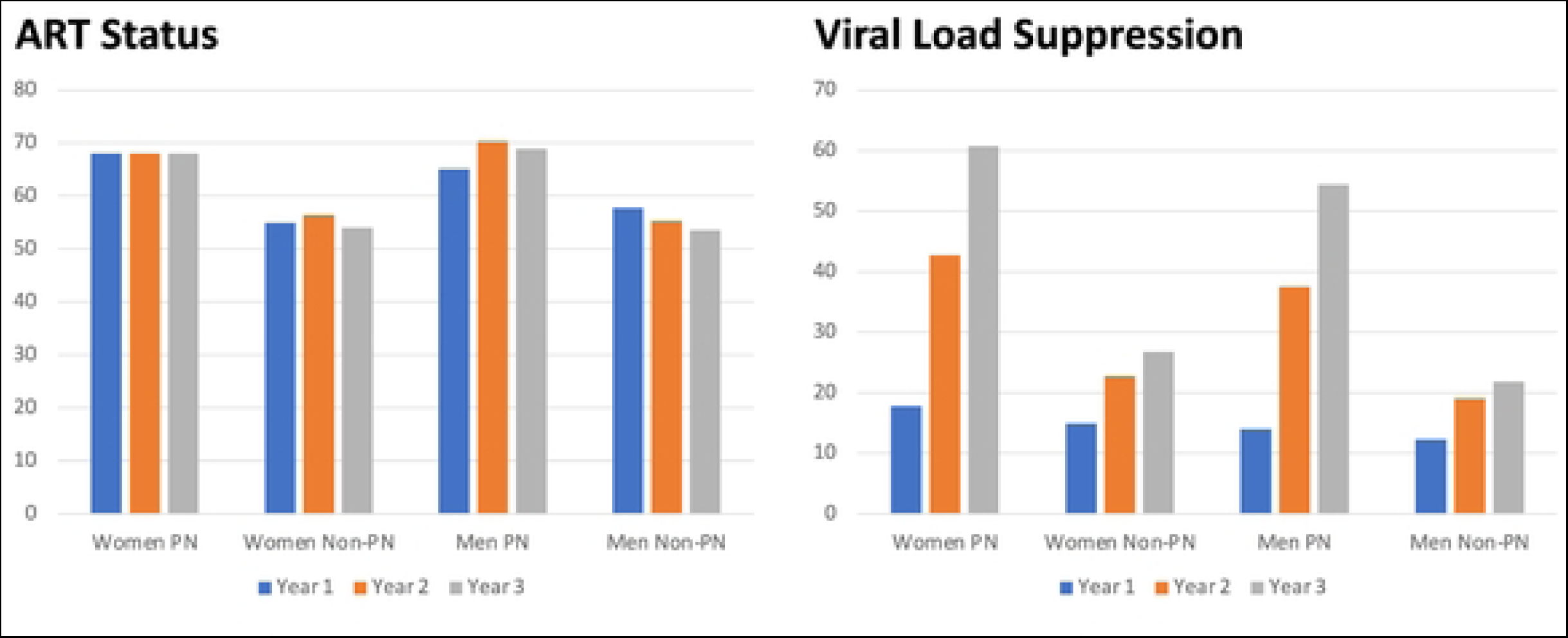
ABM Simulation of Peer Navigator Program.

The estimates of the effects of the Peer Navigator program on the population of age-specific labor force, relative to the baseline produced by SPECTRUM, are shown in Fig 4. These estimates are very small—ranging from less than 500 lives per year at the start of the forecast period to about 2,500 lives per year in 2030. The translation of these population gains into labor force gains using age and sex specific labor force participation rates from the ILO are shown in Fig 5. Finally, the impact on Tanzanian GDP is shown in Fig 6. Of course, given the small impact of the Peer Navigator program on population gains, the impacts on labor force size and GDP were correspondingly small. Nevertheless, the exercise demonstrates the feasibility of linking a hybrid ABM model that simulates the impact of a policy intervention on individual and household health seeking behaviors, translating those impacts into demographic changes, and, finally, translating the demographic changes into macroeconomic outputs.

**Fig 4.**
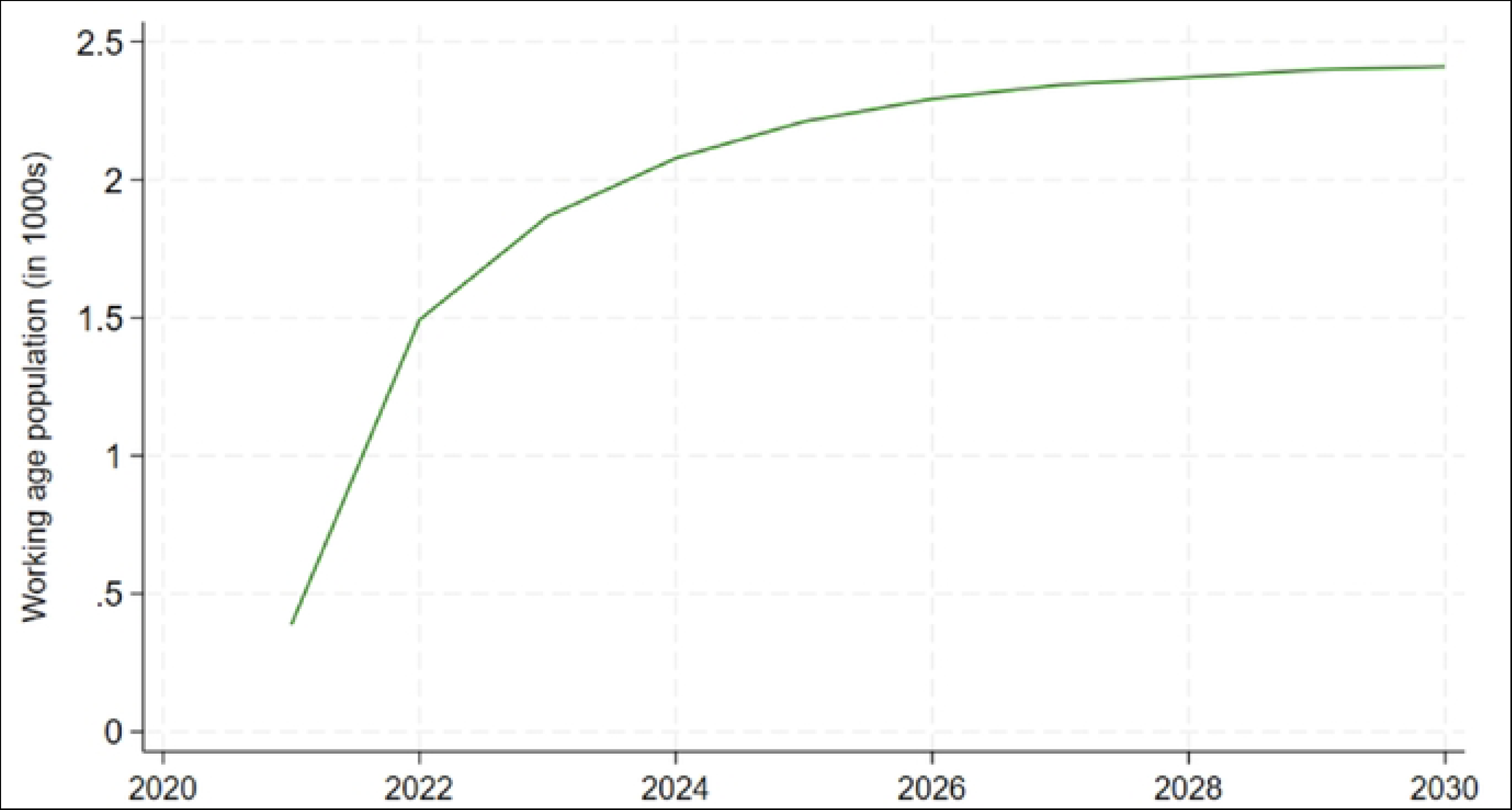
Change in Tanzania Working Age Population Resulting from the Peer Navigator Program Estimated by the SPECTRUM Model.

**Fig 5.**
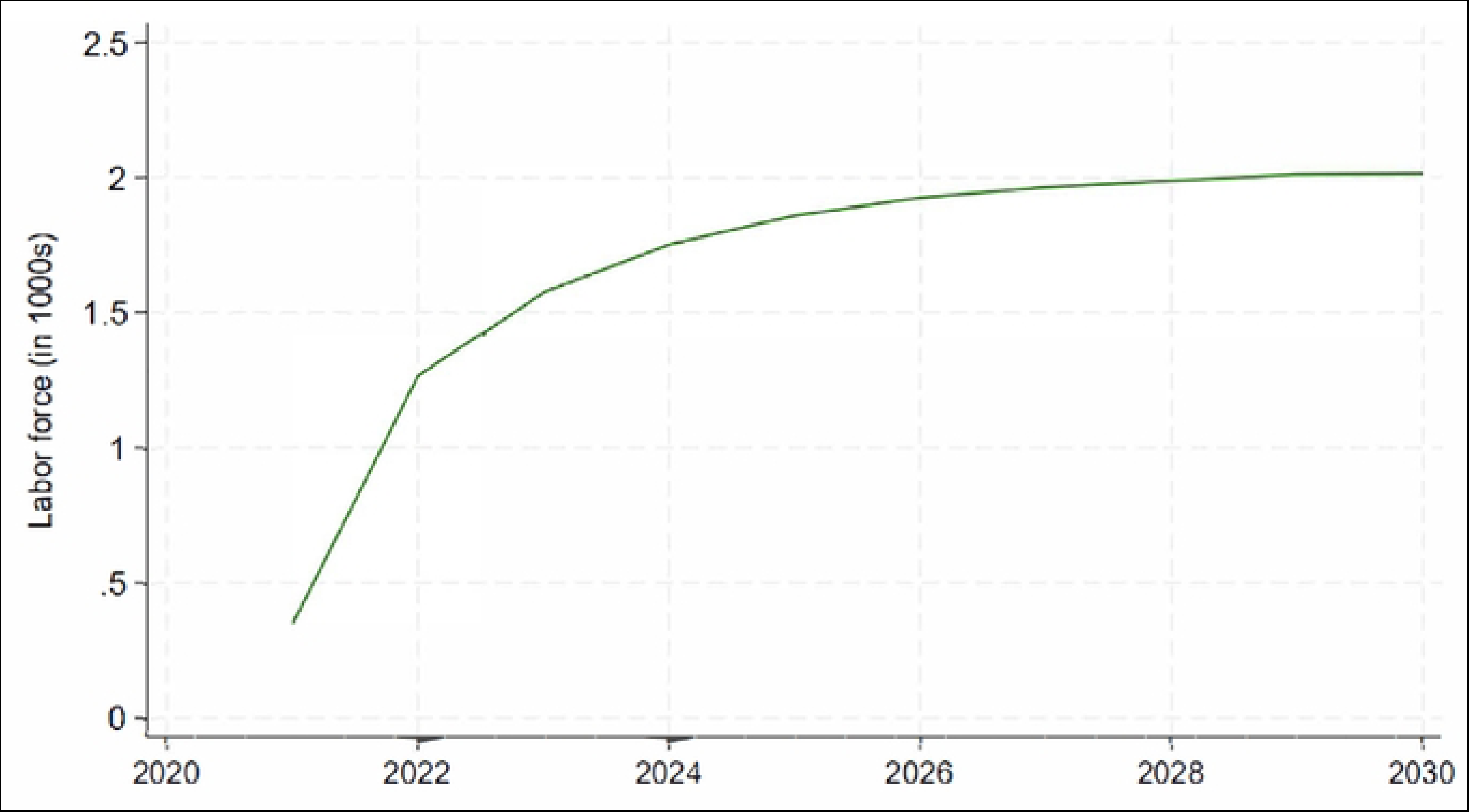
Labor Force Gains Resulting from the Peer Navigator Program.

**Fig 6.**
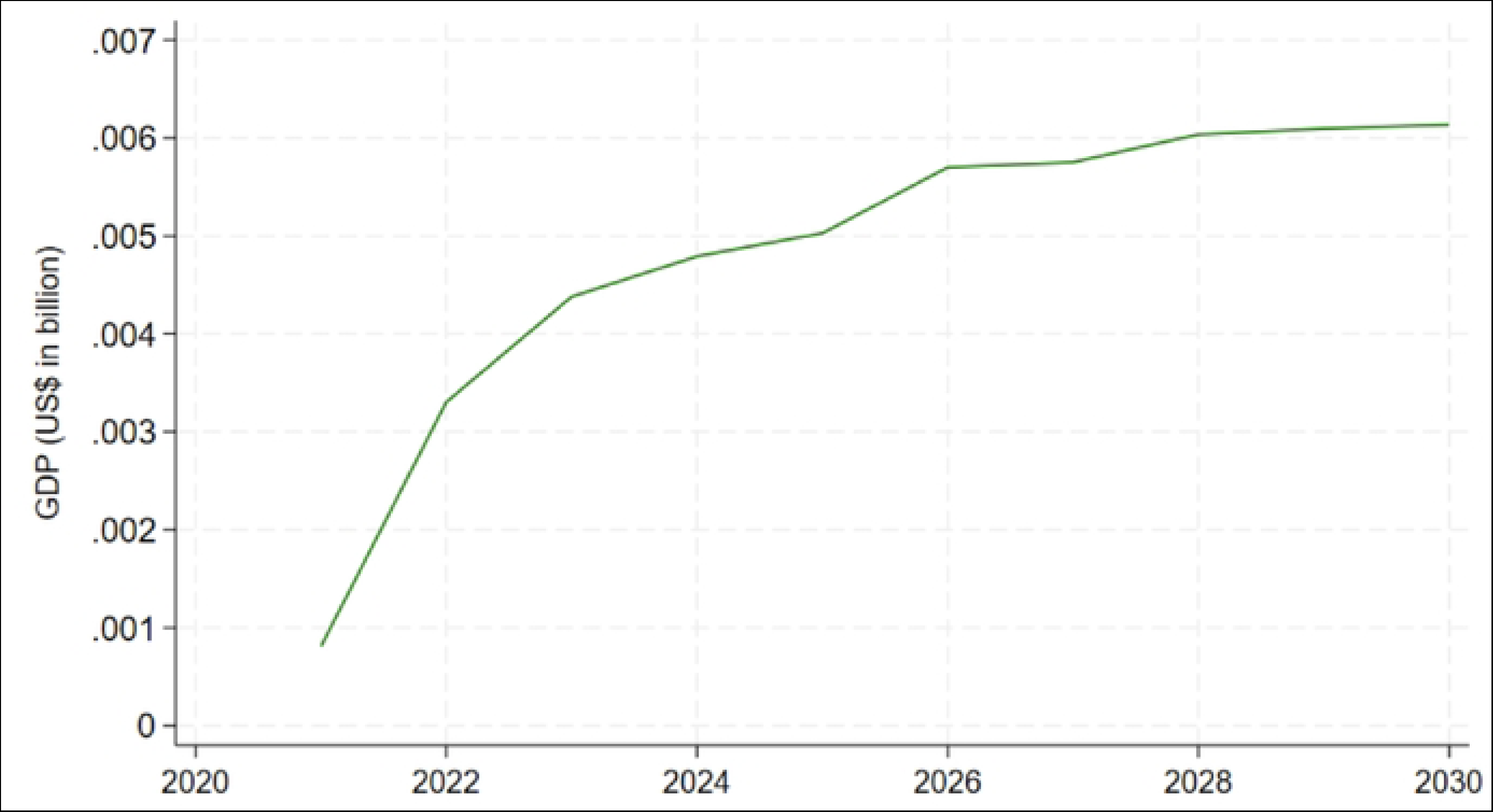
GDP Gains Resulting from the Peer Navigator Program.

## Conclusion

To our knowledge, this study represents the first successful linkage of a behavioral ABM to simulate the effects of a health policy intervention to its demographic implications and subsequent macroeconomic ramifications. Although the simulated effects of Peer Navigator Program were small, the study demonstrates the feasibility of a general approach for simulating the effects of future policy interventions with potentially larger impacts.

Although the ABM estimated that the program was highly effective in improving ART status and HIV VLS, it applied to a relatively small group of people—those in the networks of high-agency married women. In addition, Tanzania is a country that has already met its 95/95/95 goals; however, women are driving the gains in HIV care continuity. Disaggregating the overall trend in Tanzania by age and sex indicates that women outpace men and children by an average of 5 and 25 percentage points across the care cascade, respectively. (Economist Impact, 2023) While women’s agency and social capital may provide a leverage point to achieve gap closure for men and children, the concept of diminishing marginal returns implies that the opportunities for further gains in ART coverage and VLS are limited in Tanzania. The estimated modest gain in health outcomes, results in a limited increase in population size and hence macroeconomic performance. This is because only the relatively small segment of the population that have not already met the 95/95/95 goals can benefit from the incremental interventions.

### Model Limitations and Next steps

This study has demonstrated the feasibility of linking an ABM policy simulation to macroeconomic impacts through the SPECTRUM health and demographic model. However, the model would benefit from further refinements in multiple fronts. The current version of the model assumes that the age- and sex-specific labor force participation rates remain at their 2020 values over the forecast period until 2030. Similarly, trends in urbanization rates and school attendance rates are fixed at their 2020 levels. Given the slow rate of change in socio-demographic variables measured at the population level, and the fact that the forecast period extends only to 2030, we do not anticipate significant changes in the estimated macroeconomic indicators under more realistic scenarios with time-varying rates. Nevertheless, it would be preferable to replace these assumptions with estimates that recognize the endogeneity of labor force participation rates, urbanization, education, and GDP over time. This could be done by estimating an econometric model that incorporates endogeneity of labor force participation, urbanization, education, and GDP over time. Perhaps more important, the macroeconomic projects do not yet incorporate demand-side impacts resulting from rising incomes and multiplier effects as a result of health investment and rising educational levels. These effects are likely sizable over longer forecast periods. Due to underestimation of demand side stimuli, the current conservative GDP projections produced by the model should be considered lower bound estimates that mainly reflect supply-side impacts of health investment on the productive capacity of economies and only through increases in labor force participation.

## Data Availability

Data on labor force participation rates in Tanzania are available from the International Labour Organization at https://ilostat.ilo.org. Data on Tanzania Gross Domestic Product are available from the World Bank at https://datatopics.worldbank.org/world-development-indicators PHIA data used for the ABM are available from Columbia University at https://phia.icap.columbia.edu The Spectrum model used for demographic forecasts is freely downloadable at https://avenirhealth.org

https://datatopics.worldbank.org/world-development-indicators

https://phia.icap.columbia.edu

https://avenirhealth.org

## Notes

### Competing Interest Statement

The authors have declared no competing interest.

### Funding Statement

Yes

### Author Declarations

The research involved the analysis of summary-level distributions of demographic groups, labor force participation rates, and GDP. Hence, no IRB oversight or approval was required.

## References

Abuelezam NN, Rough K, & Seage III GR. (2013). Individual-Based Simulation Models of HIV Transmission: Reporting Quality and Recommendations PLoS ONE, 8(9), e75624. doi:doi:10.1371/journal.pone.0075624

Bloom D, Kuhn M, & Prettne K. (2018). IZA DP No. 11939, Health and Economic Growth. IZA Institute of Labor Economics https://www.iza.org/publications/dp/11939/health-and-economic-growth

Bloom DE, Khoury A, Kufenko V, & Prettner K. (2020). Spurring Economic Growth through Human Development: Research Results and Guidance for Policymakers. IZA Discussion Papers, No. 12964, Institute of Labor Economics: Bonn. https://www.econstor.eu/bitstream/10419/215360/1/dp12964.pdf

Bor J, Tanser F, Newell M-L, & Bärnighausen T. (2012). In a study of a population cohort in South Africa, HIV patients on antiretrovirals had nearly full recovery of employment. Health Aff (Millwood), 31(7), 1459–1469. doi:doi: 10.1377/hlthaff.2012.0407

Collin M, & Weil D. (2018). The Effect of Increasing Human Capital Investment on Economic Growth and Poverty: A Simulation Exercise, World Bank, WPS8590. https://documents1.worldbank.org/curated/en/786861537902769850/pdf/WPS8590.pdf

Crane GJ, Kymes SM, Hiller JE, &, et al. (2013). Accounting for costs, QALYs, and capacity constraints: using discrete-event simulation to evaluate alternative service delivery and organizational scenarios for hospital based glaucoma services. . Med Decis Making, 33, 986–997.

Crown W, Hariharan D, Kates J, Gaumer G, Jordan M, Hurley C, … Nandakumar A. (2023). Analysis of economic and educational spillover effects in PEPFAR countries. PLoS ONE, 18(12), e0289909. 10.1371/journal.pone.0289909

Economist Impact. 2023. A Triple Dividend: The Health, Social, and Economic Gains from Financing the HIV Response in Africa. https://www.unaids.org/en/resources/documents/2023/a-triple-dividend

Gaumer G, Luan Y, Hariharan D, Crown W, Kates J, Jordan M, … Nandakumar A. (2024). Assessing the impact of the president’s emergency plan for AIDS relief on all-cause mortality PLOS Glob Public Health, 4(1), e0002467. doi:

Gaumer G, Crown W, Kates J, Luan Y, Hariharan D, Jordan M, Hurley C, Nandakumar A. Analysis of maternal and child health spillover effects in PEPFAR countries. BMJ Open 2023;13:e070221. doi:10.1136/bmjopen-2022-070221

Geng EH, Odeny TA, Montoya LM, Iguna S, Kulzer JL, Adhiambo HF, … et al,. (2023). Adaptive strategies for retention in care among persons living with HIV. NEJM Evidence. 2(4), EVIDoa2200076.

Guo Y, Li X, & Sherr L. (2012). The impact of HIV/AIDS on children’s educational outcome: A critical review of global literature. AIDS Care, 24(8), 993–1012. doi: DOI: 10.1080/09540121.2012.668170

Homer JB, & Hirsch GB. (2006). System dynamics modeling for public health: background and opportunities. Am J Public Health, 96, 452–458.

International Labour Organization. (2023). Labour Force Statistics (LFS, LTLFS, RURBAN Databases) ILOSTAT Database Description. https://ilostat.ilo.org/resources/concepts-and-definitions/description-labour-force-statistics/

Liu P, & Wu S. (2014). An agent-based simulation model to study accountable care organizations. Health Care Manag Sci 19(1), 89–101. doi:doi: 10.1007/s10729-014-9279-x

Marshall D., Ijzerman M., Crown W., Burgos-Liz L., Osgood, Padula W., … Pasupathy K. (2015). Applying Dynamic Simulation Modeling Methods in Health Care Delivery Research: The SIMULATE checklist. Value in Health, 18(1), 5–16.

Piabuo S, & Tieguhong J. (2017). Health expenditure and economic growth - A review of the literature and an analysis between the economic community for central African states (CEMAC) and selected African countries. Health Econ Rev, 7, 23. doi:https://healtheconomicsreview.biomedcentral.com/articles/10.1186/s13561-017-0159-1

Remes J, Wilson M, & Ramdoral A. (2020). How Investing in Health Has a Significant Economic Payoff for Developing Countries. Brookings Institute. https://www.brookings.edu/blog/future-development/2020/07/21/how-investing-in-health-has-a-significant-economic-payoff-for-developing-economies/

Resch S, Korenromp E, Stover J, Blakley M, Krubiner C, Thorien K, …et al. (2011). Economic returns to investment in AIDS treatment in low and middle income countries. PLoS ONE, 6(10), e25310.

Seila AF, & Brailsford S. (2009). Opportunities and challenges in health care simulation. In Alexopoulos C, Goldsman D, & Wilson JR (Eds.), Advancing the Frontiers of Simulation. New York, NY: Springer.

Sherafat-Kazemzadeh, R., Gaumer, G., Crown, W., Daniels, E., Brown, J., Newaz, F., … Nandakumar, A. (2021). Lack of agency and sexual behaviors among married women: a study of population-based HIV impact assessment (PHIA) surveys in Malawi, Tanzania and Zambia. Journal of Global Health Reports, 5, e2021078.

Stover J, Andreev K, Slaymaker E, Gopalappa C, Sabin K, Velasquez C, …et al. (2014). Updates to the Spectrum model to estimate key HIV indicators for adults and children. AIDS, 28(Suppl 4), S427–S434.

Thirumurthy H, Galárraga O, Larson B, & Rosen S. (2012). HIV treatment produces economic returns through increased work and education, and warrants continued US support. Health Aff (Millwood), 31(7), 1470–1477.

US State Department. PEPFAR 2022 country and regional operational plan (COP/ROP) guidance for all PEPFAR-supported countries. n.d. Available: https://www.state.gov/wpcontent/uploads/2022/01/COP22-Guidance-Final_508-Compliant.pdf

Vermeer W, Gurkan C, Hjorth A, Benbow N, Mustanski BM, Kern D, …et al. (2022). Agent based model projections for reducing HIV infection among MSM: Prevention and care pathways to end the HIV epidemic in Chicago, Illinois PLoS ONE, 17(10), e0274288. doi: 10.1371/journal.pone.0274288

Vogl TS. (2012). *Education and Health in Developing Economies*. Retrieved from

Wagner Z, Borofsky J, & Sood N. (2015). PEPFAR funding associated with an increase in employment among males in ten sub-Saharan African countries. Health Affairs, 34(6). 10.1377/hlthaff.2014.1006

World Bank. (2020). Human Capital Project. https://www.worldbank.org/en/publication/human-capital

